# Population pharmacokinetic modeling of pyrotinib in patients with HER2-positive advanced or metastatic breast cancer

**DOI:** 10.1101/2020.08.23.20179655

**Authors:** Hai-ni Wen, Yi-xi Liu, Da Xu, Kai-jing Zhao, Zheng Jiao

**Author notes:** **Correspondence**: Zheng Jiao, Professor ^1^Department of Pharmacy, Shanghai Chest Hospital, Shanghai Jiao Tong University, Huaihai West Road, Shanghai, China, 200030. Hai-ni Wen and Yi-xi Liu has made equal contributions to this work.

## Abstract

**Objective:** Pyrotinib, a novel oral irreversible dual pan-ErbB tyrosine kinase inhibitor (TKI), has been approved in China for the treatment of HER2-positive advanced or metastatic breast cancer. This study aimed to conduct a population pharmacokinetics (PK) analysis of pyrotinib and to evaluate the impact of certain HER2-positive breast cancer patient characteristics on pyrotinib’s PK.

**Method:** A total of 1152 samples, provided by 59 adult female patients from two phase I clinical trials, were analyzed by nonlinear mixed-effects modeling. Monte Carlo simulation was conducted to assess the impact of covariates following exposure to pyrotinib.

**Results:** The PK of pyrotinib was adequately described by a one-compartment model with first-order absorption and elimination. Patient’s age and total protein levels can affect pyrotinib’s apparent volume of distribution, and concomitant use of montmorillonite powder had significant effects on the bioavailability of pyrotinib. No PK interactions were observed between capecitabine and pyrotinib.

**Conclusion:** In this study, a population PK model of pyrotinib was developed to determine the influence of patient characteristics on the PK of pyrotinib. While patient age and total protein levels can significantly affect the apparent distribution volume of pyrotinib, the magnitude of the impact was limited, thus no dosage adjustment was recommended. Furthermore, concomitant use of montmorillonite powder for diarrhea can decrease the bioavailability of pyrotinib by 50.3%.

## 1. Introduction

Pyrotinib is an oral, irreversible dual pan-ErbB tyrosine kinase inhibitor (TKI) that potently inhibits human epidermal growth factor receptor (HER) 1 and 2^1^. In August 2018, the National Medical Products Administration of China authorized conditional approval of pyrotinib for the treatment of HER2-positive advanced or metastatic breast cancer in patients previously treated with anthracycline or taxane chemotherapy. A dosage of 400 mg of pyrotinib once daily after a meal was recommended^2^. Furthermore, trials investigating the efficacy and safety of pyrotinib for other HER2-positive solid tumors, such as gastric cancer and non-small cell lung cancer, are on-going both in China and USA^3,4^. Moreover, according to the result of a multicentered, randomized phase II clinical trial, pyrotinib and capecitabine treatment showed greater clinical benefits than lapatinib and capecitabine treatment in patients with metastatic breast cancer. The higher overall response rate (78.5% versus 57.1%) and favored progression-free survival (18.1 versus 7.0 months) indicate that pyrotinib is a promising drug among numerous HER-family TKIs^5^.

Following oral administration of pyrotinib (80-400 mg single dose), the time required to reach maximum plasma concentration (T_max_) is 3-5 h^6,7^. Maximum concentration (C_max_) and area under the concentration-time curve (AUC) at steady state increased in a dose-dependent manner, indicating the linear pharmacokinetic (PK) profile of pyrotinib^6^. Steady-state plasma concentrations are reached within 8 days of repeated administration. No major accumulation after repeated daily administration was observed in phase I clinical trials^6^. The apparent volume of distribution (Vd/F) at steady-state was 3820 L/kg^7^. During the elimination phase, pyrotinib is mainly metabolized by the hepatic cytochrome P450 (CYP) 3A4 (>75%), and mainly excreted in the feces (>90.9%)^8,9^.

The phase I clinical trials have shown high between-subject variability following pyrotinib administration. The coefficients of variation of AUC_0-24h_ and C_max_ at steady-state varied between 25-110% and 32.8-91%, respectively^6,7^. Moreover, *in vivo* and *in vitro* studies have suggested high plasma protein binding rate (86.9-99.7%) of pyrotinib, with 58.3% covalently bound to human plasma protein^1,8^.

Therefore, understanding potential factors that can affect the PK of pyrotinib in patient populations is essential for individualized regimen dosing and optimized clinical outcomes.

Population PK (popPK) approach enables a pooled PK analysis over data from trials of heterogenous designs, as well as identification of sources of PK variability within a defined population. The aim of this study was to describe the popPK of pyrotinib in patients with HER2-positive advanced or metastatic breast cancer by pool analyzing two phase I clinical trial studies. Furthermore, the impact of covariates on pyrotinib PK, including patient demographics, biochemical laboratory tests, and co-administration of other medications was assessed.

## 2 Methods

### 2.1 Study Design and Patients

The popPK analysis was conducted using data from two phase I, single-arm, open-label, pyrotinib dose-escalation clinical trial studies. In the BLTN-Ib (NCT01937689) study, patients with HER2-positive advanced breast cancer were treated with pyrotinib monotherapy, with dosage ranging between 80 and 480 mg once daily, in 28-day cycles^7^. In the BLTN-Ic (NCT02361112) study, patients with HER2-positive metastatic breast cancer were treated with pyrotinib in combination with capecitabine. The pyrotinib dosage ranged between 160 and 400 mg, which was administered orally once daily, and capecitabine 1,000 mg/m^2^ was administered twice daily was from day 1 to 14 in 21-day cycles^^6^^. Intensive sampling was conducted in the first cycle of both regimens. A detailed description of the study design and PK sampling time points is summarized in Table 1.

**Table 1.**
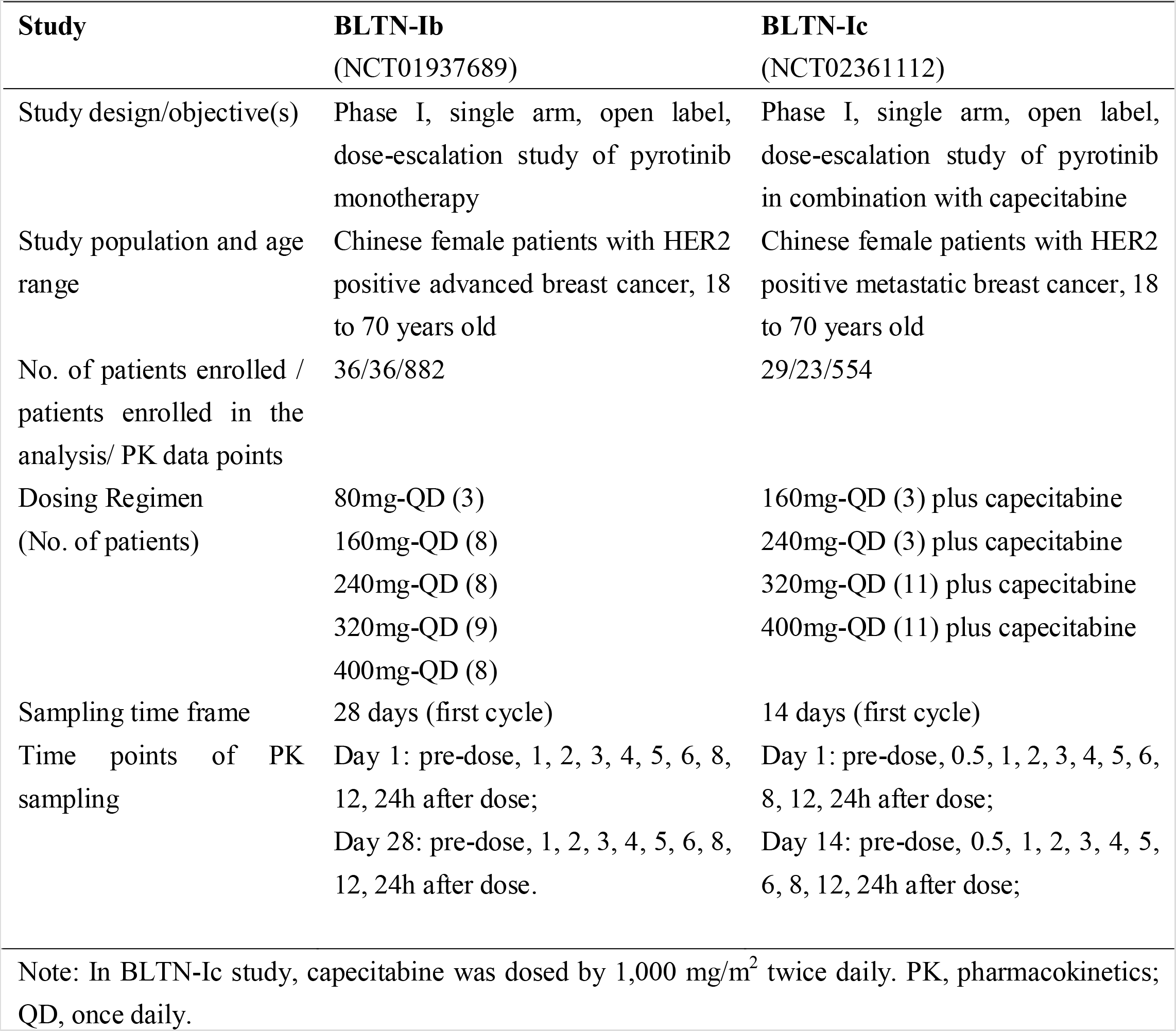
Study designs and pharmacokinetic sampling strategies for the investigated population

Both studies were conducted in accordance with the principles of the Declaration of Helsinki (October 2013) and the International Conference on Harmonization Guidelines for Good Clinical Practice^10^. Study protocols were approved by the ethics committee at each trial center. Informed consent was obtained from each participant included in each study^6,7^.

### 2.2 Bioanalytical Methods

The plasma samples were stored at –70 °C until analysis. Pyrotinib plasma concentrations were measured using a validated liquid chromatography mass spectrometry (LC/MS/MS) method. The lower limit of quantitation (LLOQ) was 0.429 ng/mL. The calibration range was 0.429 (LLOQ) to 215 ng/mL^6,7^.

### 2.3 Data handling

Patients were included for the PK analysis if they had ≥1 adequate dose and ≥1 corresponding pyrotinib plasma concentration result. Concentrations below the LLOQ were excluded from the analysis. Through exploratory analysis, data with more than six conditional weighted residual errors (CWRES) were considered potential outliers and excluded from modelling analysis. Missing values were handled as follows: 1) missing drug concentrations were documented and excluded from the analysis; 2) PK data without a sampling time were excluded from the analysis; 3) covariates with data missing for more than 10% of the patients were not included in the analysis.

### 2.4 Population Pharmacokinetic Analysis

A nonlinear mixed-effect modeling (NONMEM) approach was adopted to establish population PK models, as implemented in the NONMEM software (version 7.4.0, ICON Development Solutions, Ellicott City, MD, USA) with first-order conditional estimation with the η–x03B5; interaction (FOCE-I) method. Perl-speaks-NONMEM (PsN, version 4.7.0, Department of Pharmaceutical Biosciences, Uppsala University, Sweden) was used to perform visual predictive checks, covariate screening, and bootstrap analyses. SAS (version 9.4, SAS institute Inc, Cary, USA), R (version 3.4.1, R Foundation for Statistical Computing, Vienna, Austria) and R package “Xpose” (version 4.5.3, Department of Pharmaceutical Biosciences, Uppsala University, Sweden) were used for graphics analysis and data manipulation.

#### 2.4.1 Base Model

Based on graphical exploratory analysis, one- and two-compartment models were selected as candidate structural models. Between-subject variability (BSV) was added as a structural pharmacokinetic parameter. BSV was applied based on an exponential model, as follows:

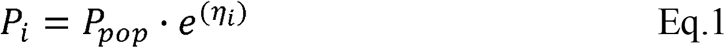

where *P_i_* stands for the individual parameter estimate for individual *i*; *P_pop_* stands for the typical population parameter estimate; *η_i_* depicts between individual random effects for individual *i*, which is assumed to adhere to a normal distribution with a mean of zero and variance of ω^2^.

Residual unexplained variability (RUV) was modelled as a proportional (Eq. 2), additive (Eq. 3) or a combination of a proportional error model and an additive error (Eq. 4) model:

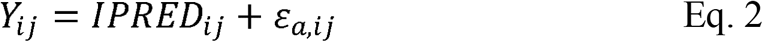

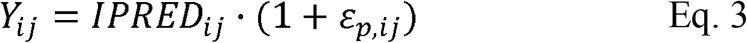

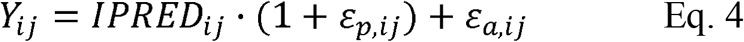

where *Y_ij_* is the observed concentration for the individual *i* at time *t_j_*; *IPRED_ij_* is the individual predicted concentration; *ɛ_p,ij_*, stands for the proportional error component, and *ɛ_a,ij_* stands for the additive error component. Residual error was assumed to adhere to a normal distribution with a mean of zero and variance of σ^2^.

Based upon Akaike’s Information Criterion (AIC)^11^, as well as the precision of parameter estimates and visual inspection of goodness-of-fit (GOF) plots, the base model was constructed.

#### 2.4.2 Covariate Model

Potential PK covariates were selected based on physiological and clinical relevance and included: age, total body weight (BW), body mass index (BMI), albumin, globulin, total protein (TP), aspartate aminotransferase (AST), alanine aminotransferase (ALT), total bilirubin (TB), serum creatinine (SCr), estimated glomerular filtration rate (eGFR), and occurrence of diarrhea. Co-administered medications were included in covariate analysis if more than 10% of patients received the medication.

All candidate covariates were first graphically explored to identify potential correlations and then carried through to subsequent analyses based on mechanistic plausibility and clinical relevance. Highly correlated covariates were identified through the graphical analysis and only one of the correlated covariates was used for further analysis.

Subsequently, potential covariates were examined using stepwise covariate modeling method, including the forward inclusion and backward elimination process. During the forward inclusion process, the potential covariates were added to the base model one at a time, and the addition of one parameter required a significant decrease in objective function value (>3.84, *p* < 0.05). A complete model was obtained when all significant covariates were included. During the backward elimination process, covariates from the complete model were removed one at a time, and the criterion for retention of a covariate was a significant change in objective function value (>10.83, *p* < 0.001) following the elimination.

Continuous covariates were assessed by both a linear function and a power function (Eq. 5 and 6), while categorical covariates were tested with Eq. 7:

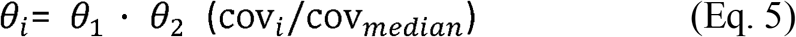

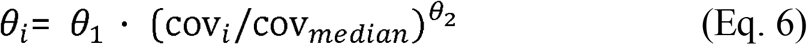

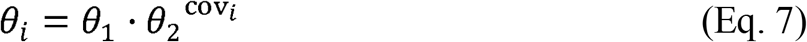

where *θ_i_* and *cov_i_* describe the parameter and covariate value for the individual *i*, respectively. cov is the median value for each covariate. *θ*_1_ represents the typical value of a pharmacokinetic parameter at the median covariate value, and *θ*_2_ is an estimated value. In Eq. 7, *cov_i_* equals to 0 or 1 for categorial variables.

#### 2.4.3 Model evaluation

Model evaluation was performed by using GOF plots for model fitness. Scatterplots were used to evaluate the observed values *versus* population predicted values, observed values *versus* individual predicted values, CWRES *versus* population predicted values, and CWRES *versus* time. In addition, nonparametric bootstrap resampling (n = 2000) technique was employed for model evaluation. Each parameter was calculated repeatedly by applying the final model to 2000 bootstrapped datasets. The 2.5th and 97.5th percentiles and the median of the popPK parameter estimates from the resampled datasets were compared with the estimates of the final model.

In order to adjust for differences of independent variables, a prediction corrected visual predictive check (pcVPC) method was employed for simulation-based model assessment^12^. The pcVPC approach was conducted by simulating 1000 datasets using the parameters of the final model, and by comparing the 5th and 95th percentiles and the median in the observed data with the corresponding percentiles and median in the simulated data. The concordance of variability and central tendency between the observed and simulated data was evaluated.

#### 2.4.4 Model-based simulation

Monte Carlo simulations were employed to evaluate the effect of covariates on the PK of pyrotinib following 400 mg oral administration once daily. PK profile of a typical reference patient (age 47 years, total protein 72.8 g/L) was compared to that of patients with various covariate levels, which were set as 10th and 90th percentile of population distribution. The predicted pyrotinib exposures were based on 1000 simulations for each group of patients, applying final model parameters for both fixed and random effects.

## 3. Result

### 3.1 Patient characteristics and pharmacokinetic datasets

Out of 1436 PK data points from the original dataset, the final dataset contained a total of 1152 (80.22%) plasma samples, provided by 59 (19.5 samples per person) adult patients from BLTN-Ib (36 patients, 670 samples) and BLTN-Ic study (23 patients, 482 samples). According to prespecified exclusion criteria, 284 concentrations were excluded from the analysis: there were 79 observations (42 from BLTN-Ib study and 37 from BLTN-Ic study) below LLOQ; records of sampling time were missing for 205 data points (170 samples from BLTN-Ib and 35 from BLTN-Ic). No data were recognized as outliers.

Patient demographics, laboratory measurements, and co-administered medications are summarized in Table 2. The female adult patients diagnosed with HER2-positive advanced or metastatic breast cancer enrolled in the PK analysis had a median age of 47 years old and a median body weight of 61 kg. More than 90% of the participants had total protein levels within the reference range (60–83 g/L) and most patients had normal renal and hepatic functions. Interestingly, the occurrence of diarrhea was much higher in patients from the BLTN-Ic group (19.60%) than that of the BLTN-Ib group (5.68%). The patients from the BLTN-Ic group were administered montmorillonite powder for diarrhea treatment. Overall, 68 out of 115 (59.13%) PK data points were sampled from patients receiving montmorillonite powder for diarrhea treatment. In addition, patients with concomitant use of capecitabine and pyrotinib were all from the BLTN-Ic group.

**Table 2.**
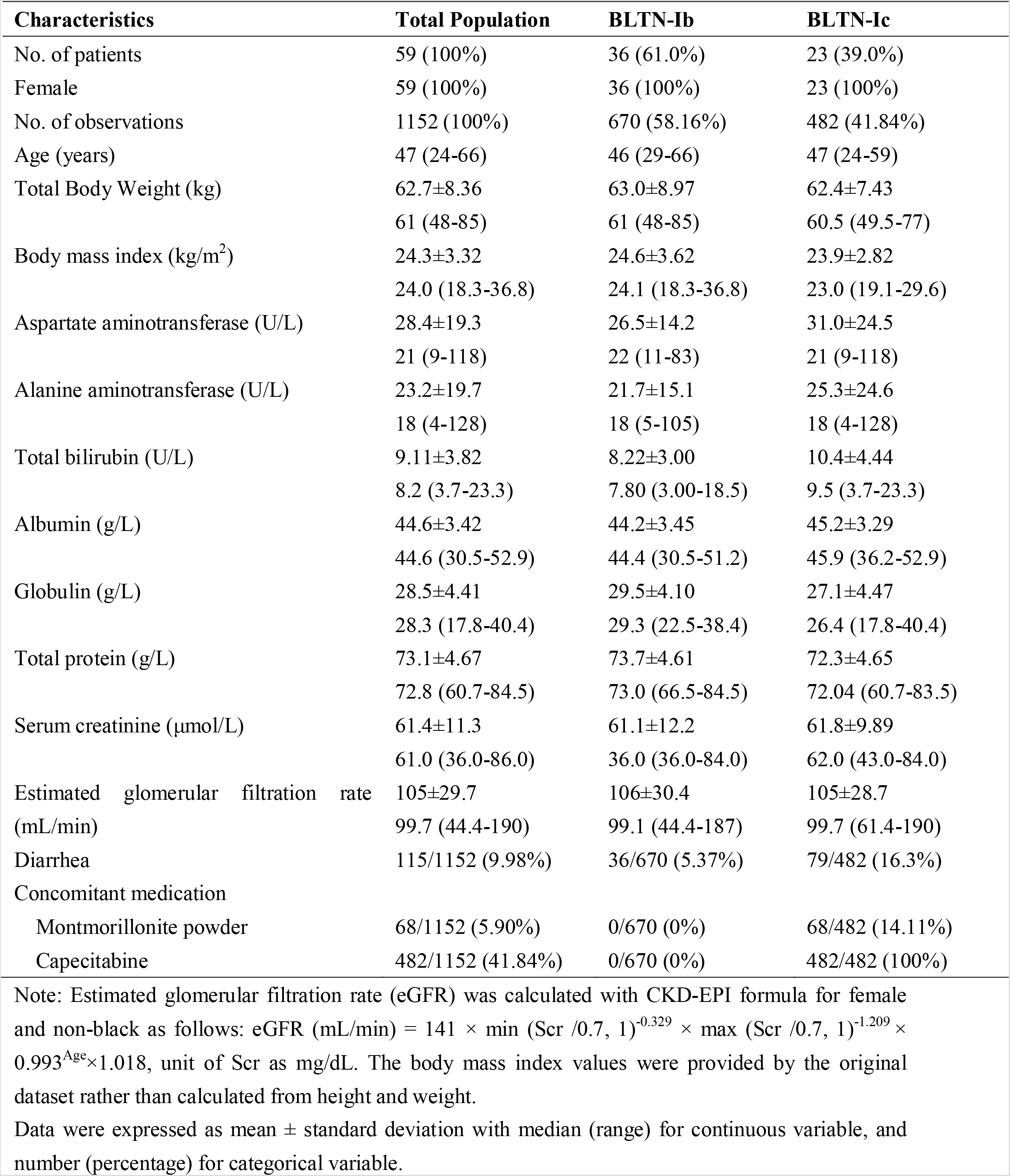
Baseline demographics and disease characteristics of included patients.

### 3.2 Population Pharmacokinetic Model

#### 3.2.1 Base Model

PK data from the two studies were best described using a one-compartment model with first-order absorption and elimination. The adoption of a one-compartment model showed increased precision of parameters and decreased Akaike’s information criterion by 306.7, compared to the two-compartment model. The model was parameterized by apparent clearance (CL/F), Vd/F, and absorption rate constant (*k*_a_). The BSV was incorporated into all parameters. A combination of proportional and additive error model was adopted for RUV in BLTN-Ib and BLTN-Ic studies respectively, considering that PK sampling time as well as concomitant medications were different in the two studies.

#### 3.2.2 Covariate Model

Graphical exploratory analysis revealed that the BMI was correlated with BW; albumin and globulin were correlated with TP; ALT was correlated with AST; SCr was correlated with eGFR. Therefore, BW, AST, TP, and eGFR were selected as covariate candidates due to their clinical relevance. Based on visual examination, the effects of age, TB, and AST were evaluated as covariates of CL/F, while the effects of age, BW, and TP were evaluated as covariates of Vd/F. The co-administration of pyrotinib with montmorillonite powder or capecitabine were evaluated as covariates of all parameters. Occurrence of diarrhea was evaluated as a covariate of *k*_a_ and bioavailability.

During the forward inclusion step, the effects of TP, age, AST, co-administration of capecitabine, and co-administration of montmorillonite powder were included, while AST and co-administration of capecitabine were eliminated in the subsequent backward elimination step. The major steps in the base and covariate model building are provided in Supplementary Material (Table S1).

The final model is listed below:

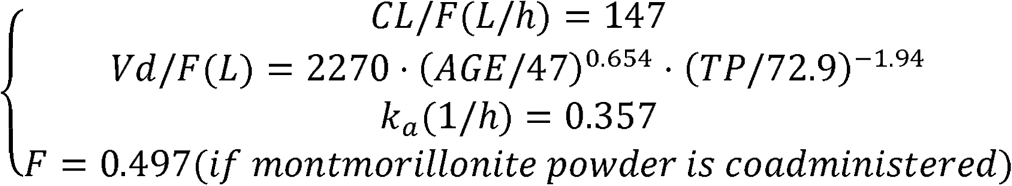

The parameters of the final model are shown in Table 3. All shrinkages of BSV and RUV were less than 30%, suggesting a reliable estimate of each parameter. Of noted, the inclusion of TP and age as covariates of Vd/F has led to a decrease of BSV from 45.72% to 40.12%, indicating that 5.6% of BSV in Vd/F was explained by TP and age. Co-administration of montmorillonite powder for diarrhea treatment can decrease the bioavailability of pyrotinib by 50.3%. Co-administration of capecitabine showed no effects on the PK of pyrotinib.

**Table 3.** Population pharmacokinetic parameter estimates of final model and bootstrap evaluation.

| Parameter | Final model |  | Bootstrap |  | Relative |
| --- | --- | --- | --- | --- | --- |
|  | Estimates | RSE% | Median | 2.5th-97.5th percentile | Bias (%) |
| Structural model parameter |  |  |  |  |  |
| CL | 147 | 6.40 | 147 | 129–167 | -0.08% |
| V | 2270 | 7.00 | 2276 | 1972–2593 | 0.28 |
| $k_a$ | 0.357 | 11.10 | 0.358 | 0.291–0.443 | 0.28 |
| MP on F | 0.497 | 14.50 | 0.497 | 0.338–0.619 | -0.03 |
| AGE on V | 0.654 | 21.10 | 0.652 | 0.342–0.917 | -0.28 |
| TP on V | -1.94 | 32.90 | 1.89 | 0.698–3.34 | -2.78 |
| Between-subject variability |  |  |  |  |  |
| $\omega_{CL}$ (%CV) | 50.20 | 25.10 | 48.92 | 37.5–62.3 | -2.55 |
| $\omega_V$ (%CV) | 40.12 | 23.30 | 39.08 | 29.4–48.8 | -2.59 |
| $\omega_{ka}$ (%CV) | 59.41 | 26.10 | 58.28 | 42.0–74.7 | -1.91 |
| $\omega_{CL-V}$ (%CV) | 82.90 | 25.60 | 82.50 | 81.1–84.9 | -0.48 |
| Residual unexplained variability |  |  |  |  |  |
| Proportional1 (%CV) | 25.30 | 17.00 | 24.87 | 19.4–29.6 | -1.68 |
| Additive 1 (SD) | 0.0071 | 55.20 | 0.0073 | 0.00326–0.0127 | 2.54 |
| Proportional2 (%CV) | 18.60 | 27.20 | 18.40 | 13.2–23.5 | -1.08 |
| Additive 2 (SD) | 0.0150 | 27.20 | 0.0150 | 13.2–23.5 | -0.14 |

AIC, Akaike information criterion; CL, clearance ( L/h); $k_a$ , absorption rate constant; V, volume (L); MP on F, influence of co-administration of montmorillonite powder for absolute bioavailability; AGE on V, influence of age for volume; TP on V, influence of total protein for volume; $\omega_{CL}$ , between-subject variability of clearance; $\omega_V$ , between-subject variability of volume; $\omega_{ka}$ , between-subject variability of absorption rate constant; $\omega_{CL-V}$ , covariance of clearance and volume; Proportional1,proportional error of Ib study; Additive1, additive error of Ib study; Proportional2, proportional error of Ic study; Additive2, additive error of Ic study.

#### 3.2.3 Model Evaluation

The GOF diagnostic plots of the final model (Fig. 1) showed adequate agreement between predicted and observed pyrotinib plasma concentration values, with no indication of model bias over a wide range of concentrations. Over time and across predicted pyrotinib plasma concentration values no bias was observed in CWRES, suggesting that the final model adequately described the PK of pyrotinib. The final model was resampled 2000 times by non-parametric bootstrapping, with 1984 computations successfully presenting the minimization step. The bootstrapped median and 2.5th and 97.5th percentile values of each parameter were consistent with the final model parameter estimates (Table 3).

**Figure 1.**
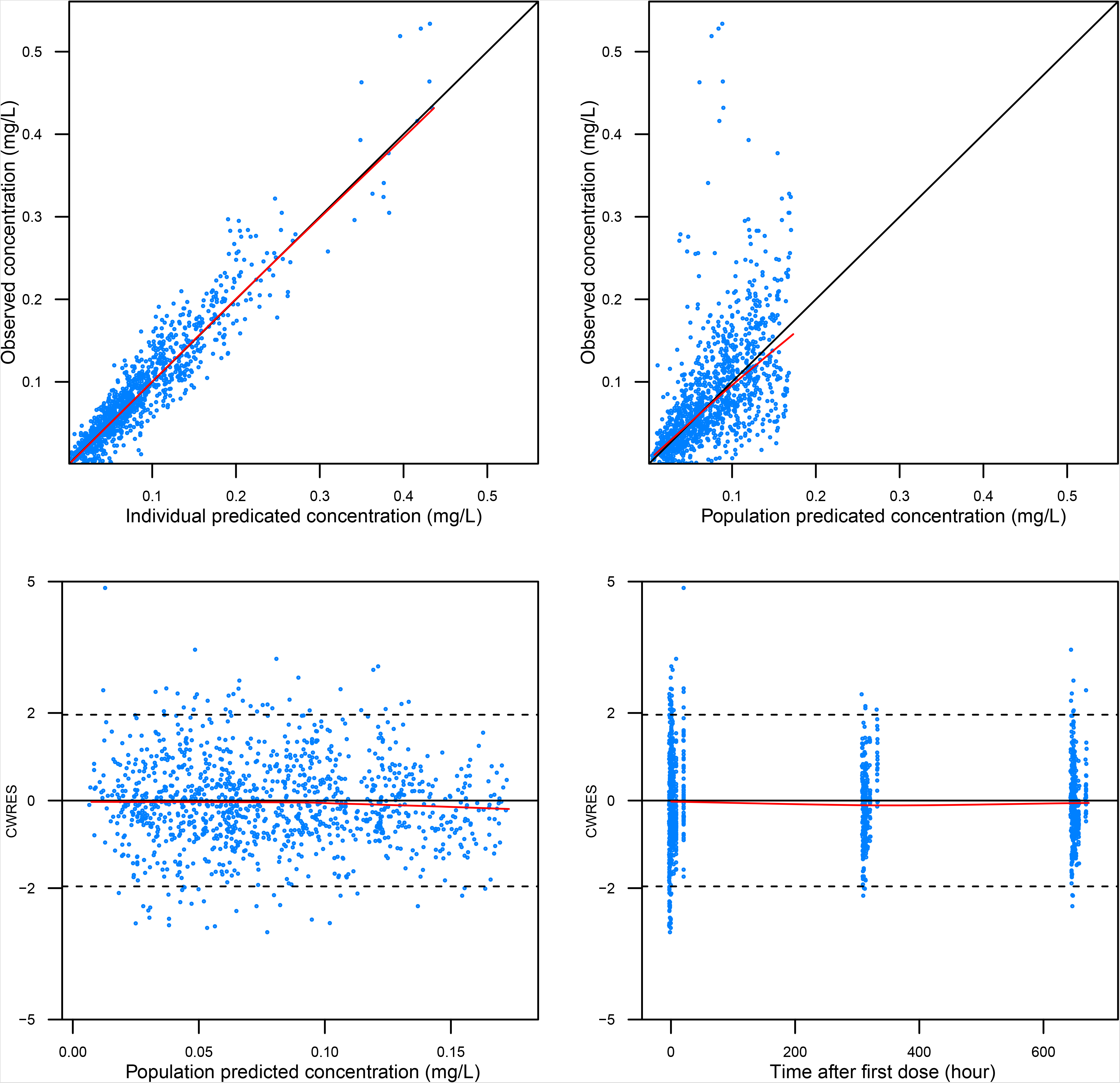
Goodness-of-fit plots of the final population-pharmacokinetic model. The red line represents the locally weighted scatterplot smoothing line.

As presented in Figure 2, the pcVPC assessment demonstrated that the final model adequately characterized the trend of the concentration-time profile. Overall, the pcVPC plots showed that the median, 5th and 95th percentile of the simulated pyrotinib concentrations largely overlapped with the observed values, indicating a reasonable predictive performance of the final model.

**Figure 2.**
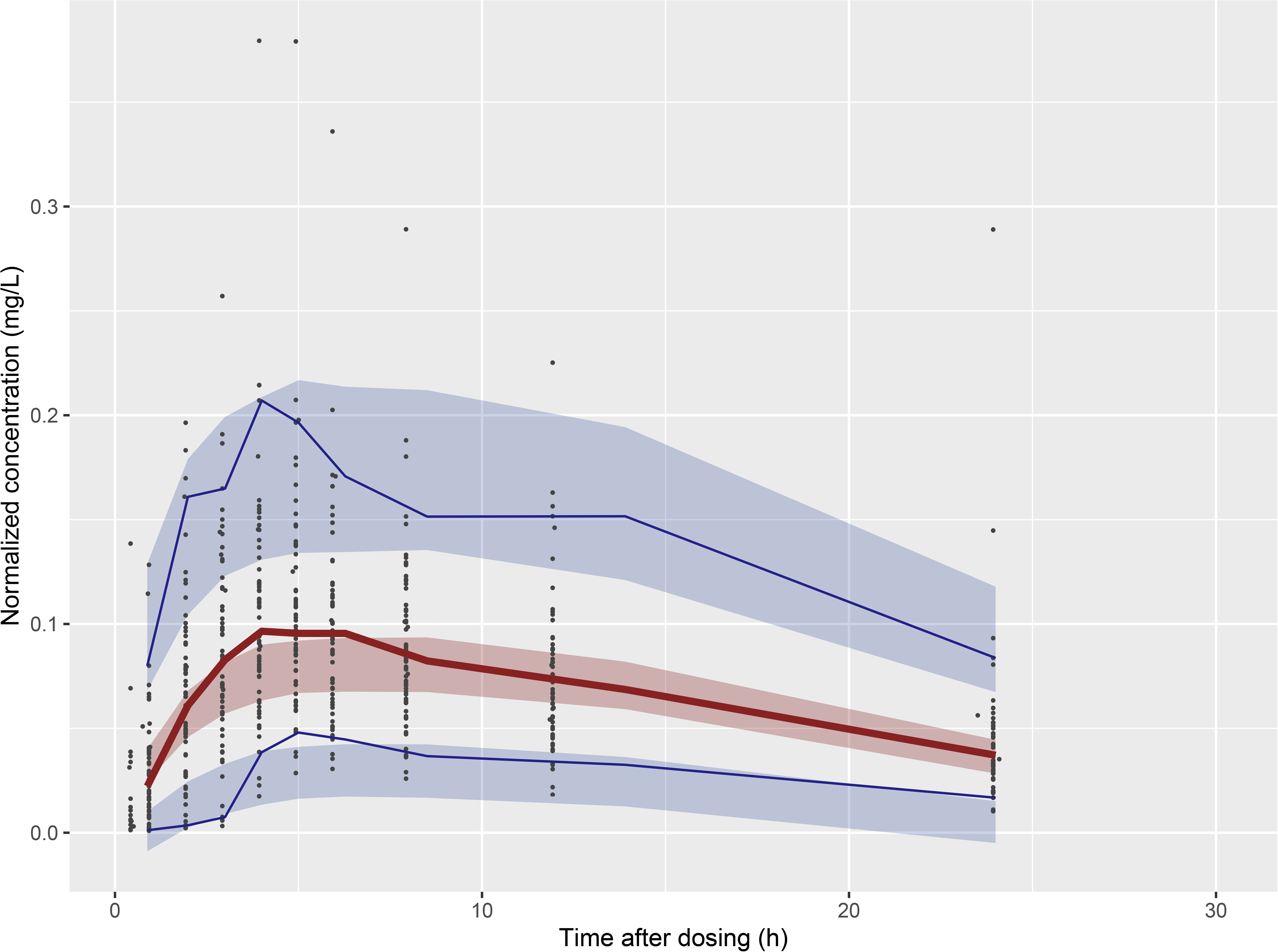
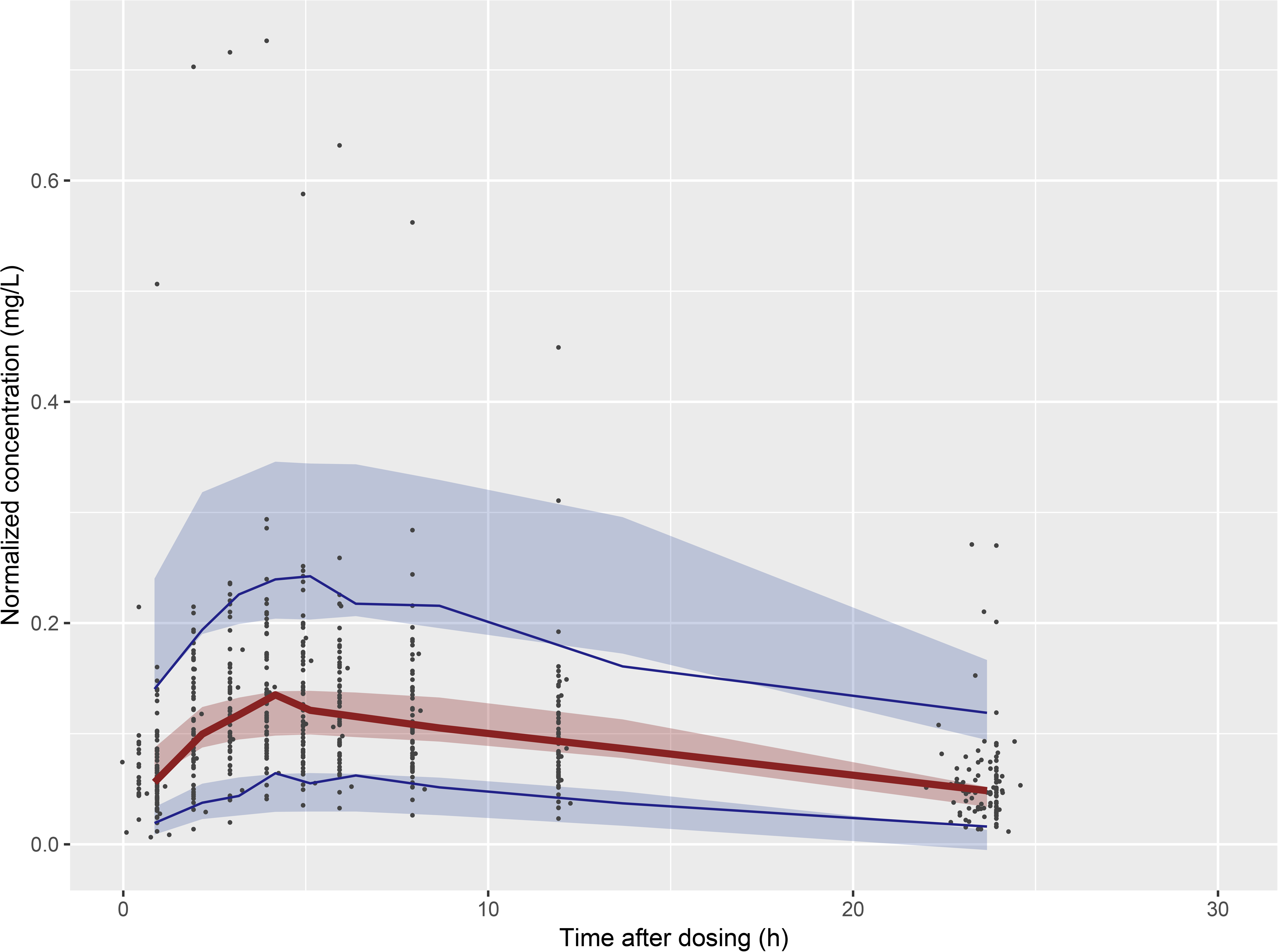
Prediction-corrected visual predictive check of the final model. Dots represent the observed data. Solid lines represent the 5th (blue), 50th (red bold), and 95th (blue) percentiles of the observed data. Shaded areas represent nonparametric 95% confidence intervals for the 5th (light blue), 50th (light red), and 95th (light blue) percentiles of the corresponding model-predicted percentiles.

#### 3.2.4 Model-based simulation

The simulated PK profiles of pyrotinib following administration of 400 mg once daily in patients of varying TP levels and ages are shown in Figure 3. Higher steady-state pyrotinib concentrations were observed in younger patients and patients with higher TP levels. Furthermore, the simulated concentration-time profiles suggested that age and TP levels had limited effects on plasma concentration of pyrotinib.

**Figure 3.**
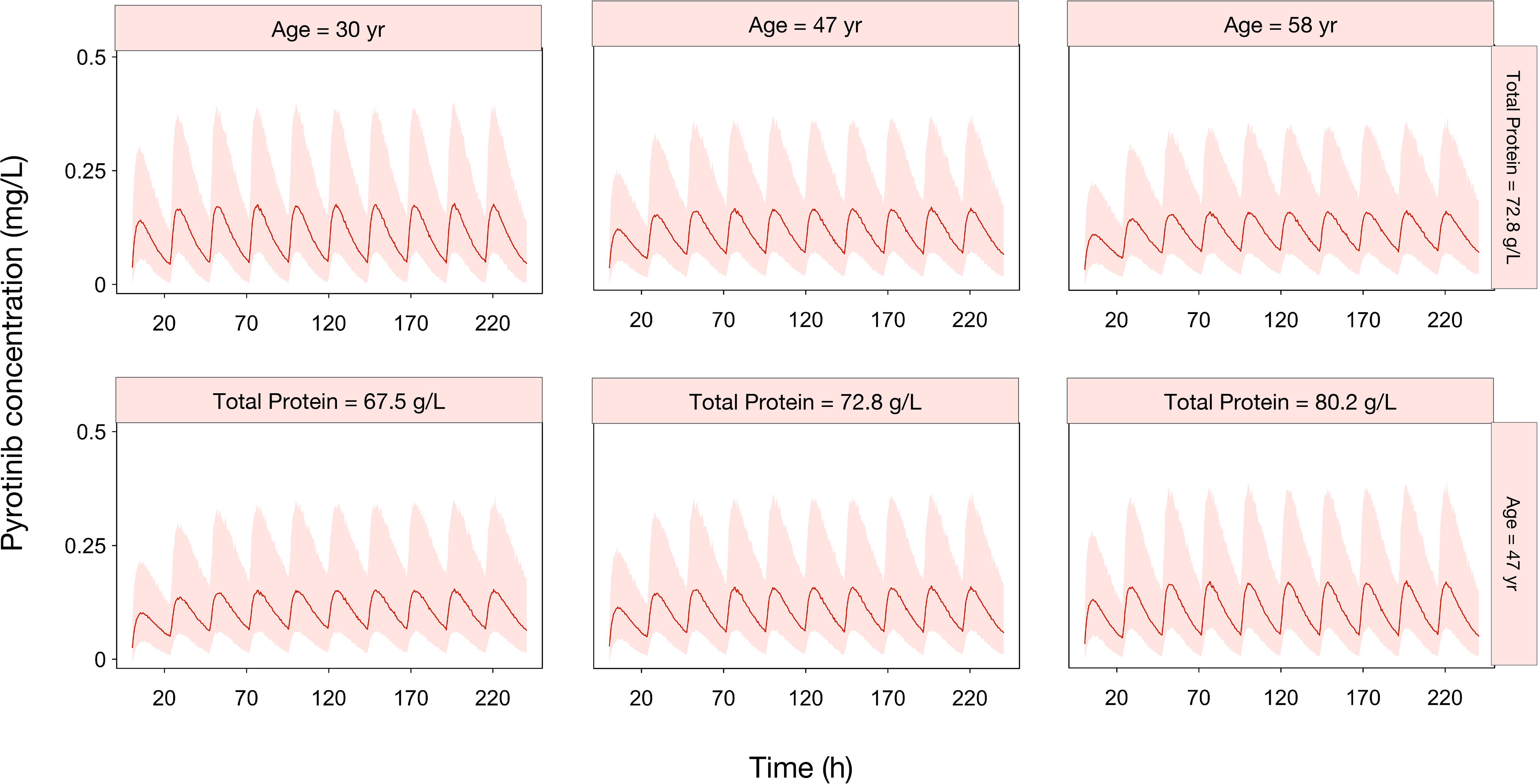
Simulated median pyrotinib plasma concentration-time curves following oral administration of 400 mg once daily. The typical reference patient was 47 years old and had total protein levels of 72.8 g/L. The solid lines represent the median values of 1000 simulations, and the shaded area is the 95% prediction interval.

## 4. Discussion

In this study, for the first time, a pyrotinib popPK model was developed, using a total of 1152 pyrotinib plasma samples from 59 adult female patients with HER2-positive advanced or metastatic breast cancer. A one-compartment model with first-order absorption and first-order elimination obtained the best description for the pyrotinib profile. The final model-derived typical values of pyrotinib PK were 147 L/h for CL/F, 2270 L for Vd/F, and 0.352 for absorption constant *k*_a_.

While age and TP levels were identified as significant covariates of pyrotinib Vd/F during popPK modelling process, the effects of both covariates appeared to be limited. Thus, dose adjustment according to TP levels or age is not necessary. Nevertheless, the current work has identified and quantified sources of inter-individual variability in pyrotinib response, which has laid the groundwork for further PK studies as well as precision dosing strategies.

Consistent with previous popPK analysis on other TKIs, the minor effect of age on the effect of pyrotinib was also shown in afatinib, erlotinib, and sunitinib, and all studies have concluded that no dose adjustment was necessary due to the low clinical relevance^13, 14, 15^. As pyrotinib is highly lipophilic (logP = 4.475)^15, 16^, the influence of age on pyrotinib distribution volume revealed in this study could be explained by age-related increase of fat and changes in adipose tissue.

The influence of TP level on Vd/F can be explained by the affinity of pyrotinib to human plasma protein. The TP represents the total amount of albumin and globulin, both of which are vital physiological components. According to previous findings, the *α, β*-unsaturated amide structure of pyrotinib molecule is highly affinitive to proteins, and able to covalently bind to human plasma protein^1,8^. Thus, 86.9–99.7% of pyrotinib was bound to plasma proteins following oral administration^2^. As a result, patients with higher protein levels would experience higher pyrotinib plasma concentration. Most patients included in our analysis had normal TP levels; therefore, whether patients with extremely low TP levels, such as those under poor nutrition status, need dosage adjustment should be further studied with a larger patient population.

Our study found that concomitant use of montmorillonite powder can decrease the bioavailability of pyrotinib by 50.3%, whereas occurrence of diarrhea failed to be included as a significant covariate affecting the pyrotinib response. Montmorillonite powder is an adsorbent commonly prescribed for diarrhea treatment^18^. Due to its unique structure as an aluminosilicate, montmorillonite can strongly adsorb to toxins, and protect the mucosal barrier, by binding mucous glycoproteins and preventing toxin absorption^19^. Therefore, it is believed that concomitant use of an adsorbent could reduce the bioavailability of pyrotinib through adsorption or decreasing its systemic absorption^20^.

Considering the possibility that the occurrence of diarrhea could also impede drug absorption in the GI tract^21^, confounding bias cannot be excluded due to the following limitations: First, the impact of non-adsorbent antidiarrheals was not investigated due to data unavailability. Second, occurrence of diarrhea was included in the analysis as a dichotomous rather than polytomous covariate due to data characteristics. Thus, the impact of diarrhea severity was not investigated. However, patients with higher severity of diarrhea have often been treated with montmorillonite powder. In addition, as shown in Table 1, more than 50% of patients who experienced diarrhea were treated with montmorillonite powder. Therefore, the current finding needs to be further supported by studies using a larger sample size.

No significant PK drug-drug interaction was observed between capecitabine and pyrotinib in our analysis. Capecitabine is a weak inhibitor of CYP2C9^22^. Pyrotinib is a major substrate of CYP3A4 (>75%) and minor substrate of CYP3A5^1^. The combination of pyrotinib and capecitabine has been approved as treatment for HER2-positive advanced breast cancer. Our study suggests that combining these drugs does not compromise the PK of either drug.

Several limitations in our study should be addressed. First, the study included only Chinese female patients with breast cancer. While pyrotinib use is expanding to other solid tumors characterized by HER2 overexpression, whether the conclusions presented here could be generalized to patients of another cancer type, or populations of different races or sexes, remains to be further investigated. Second, as the dose-response of pyrotinib has not been well-established, the applicability of the current PK model for optimizing clinical dosing strategies is limited.

## 5. Conclusion

In this study, a popPK model of pyrotinib for patients with advanced or metastatic breast cancer was developed for the first time. The concomitant use of montmorillonite powder for diarrhea treatment was shown to decrease the bioavailability of pyrotinib by 50.3%. Furthermore, no PK interaction was shown between pyrotinib and capecitabine. While pyrotinib distribution volume can be significantly affected by patient age and TP levels, the magnitude of the impact was limited, thus no dosage adjustments were recommended.

## Data Availability

Due to the nature of this research, participants of this study did not agree for their data to be shared publicly, so supporting data is not available.

## Abbreviations

ALT: alanine aminotransferase
AST: aspartate aminotransferase
AUC: concentration-time curve
BMI: body mass index
BSV: between-subject variability
BW: body weight
CL/F: apparent clearance
CYP: cytochrome P450
CWRES: conditional weighted residual error
eGFR: estimated glomerular filtration rate
GOF: goodness-of-fit
HER: human epidermal growth factor receptor
ka: absorption rate constant
LLOQ: lower limit of quantitation
pcVPC: prediction corrected visual prediction check
popPK: population PK
PK: pharmacokinetics
RUV: residual unexplained variability
SCr: serum creatinine
TB: total bilirubin
TKI: tyrosine kinase inhibitors
TP: total protein
Vd/F: apparent volume of distribution

## Acknowledgements

We thank Dr. Guang-li Ma, Dr. Yu-ya Wang, and Miss Mei-xia Chen from Jiangsu Hengrui Medicine Co. Ltd for critical reviewing of the work. We also thank Editage (www.editage.cn) for English language editing.

## CRediT author statement

**Hai-ni Wen**: Methodology, Validation, Visualization, Writing - Original Draft, Writing - Review & Editing. **Yi-xi Liu**: Methodology, Software, Formal analysis, Writing - Original Draft, Writing - Review & Editing. **Da Xu**: Data Curation, Writing - Review & Editing. **Kai-jing Zhao**: Data Curation, Writing - Review & Editing. **Zheng Jiao:** Conceptualization, Methodology, Supervision, Writing - Original Draft, Writing –Review & Editing.

## Reference

1. Li X, Yang C, Wan H, et al. Discovery and development of pyrotinib: A novel irreversible EGFR/HER2 dual tyrosine kinase inhibitor with favorable safety profiles for the treatment of breast cancer. Eur J Pharm Sci. 2017;110:51–61. doi:10.1016/j.ejps.2017.01.021

2. Blair HA. Pyrotinib: First Global Approval. Drugs. 2018;78(16):1751–1755. doi:10.1007/s40265-018-0997-0

3. Zhou C, Li X, Wang Q, et al. Pyrotinib in HER2-Mutant Advanced Lung Adenocarcinoma After Platinum-Based Chemotherapy: A Multicenter, Open-Label, Single-Arm, Phase II Study. J Clin Oncol. July 2020:JCO.20.00297. doi:10.1200/JCO.20.00297

4. Gao Z, Song C, Li G, et al. Pyrotinib treatment on HER2-positive gastric cancer cells promotes the released exosomes to enhance endothelial cell progression, which can be counteracted by apatinib. Onco Targets Ther. 2019;12:2777–2787. doi:10.2147/OTT.S194768

5. Ma F, Ouyang Q, Li W, et al. Pyrotinib or Lapatinib Combined With Capecitabine in HER2-Positive Metastatic Breast Cancer With Prior Taxanes, Anthracyclines, and/or Trastuzumab: A Randomized, Phase II Study. J Clin Oncol. 2019;37(29):2610–2619. doi:10.1200/JCO.19.00108

6. Li Q, Guan X, Chen S, et al. Safety, efficacy, and biomarker analysis of pyrotinib in combination with capecitabine in HER2-positive metastatic breast cancer patients: A phase i clinical trial. Clin Cancer Res. 2019;25(17):5212–5220. doi:10.1158/1078-0432.CCR-18-4173

7. Ma F, Li Q, Chen S, et al. Phase I study and biomarker analysis of pyrotinib, a novel irreversible Pan-ERBB receptor tyrosine kinase inhibitor, in patients with human epidermal growth factor receptor 2–positive metastatic breast cancer. J Clin Oncol. 2017;35(27):3105–3112. doi:10.1200/JCO.2016.69.6179

8. Meng J, Liu X yun, Ma S, et al. Metabolism and disposition of pyrotinib in healthy male volunteers: covalent binding with human plasma protein. Acta Pharmacol Sin. 2019;40(7):980–988. doi:10.1038/s41401-018-0176-6

9. Yunting Zhua, Liang Lia, Ge Zhang, Hong Wan, Changyong Yang, Xingxing Diao, Xiaoyan Chen, Lianshan Zhang DZ. Metabolic characterization of pyrotinib in humans by ultra-performance liquid chromatography/quadrupole time-of-flight mass spectrometry. J Chromatogr B. 2016;1033-1034:117-127. doi:10.1016/J.JCHROMB.2016.08.009

10. Malik AY, Foster C. The revised Declaration of Helsinki: cosmetic or real change? J R Soc Med. 2016;109(5):184–189. doi:10.1177/0141076816643332

11. Donohue MC, Overholser R, Xu R, Vaida F. Conditional Akaike information under generalized linear and proportional hazards mixed models. Biometrika. 2011;98(3):685–700. doi:10.1093/biomet/asr023

12. Bergstrand M, Hooker AC, Wallin JE, Karlsson MO. Prediction-corrected visual predictive checks for diagnosing nonlinear mixed-effects models. AAPS J. 2011;13(2):143–151. doi:10.1208/s12248-011-9255-z

13. Nakao, K., Kobuchi, S., Marutani, S. et al. Population pharmacokinetics of afatinib and exposure-safety relationships in Japanese patients with EGFR mutation-positive non-small cell lung cancer. Sci Rep 9, 18202 (2019). https://doi.org/10.1038/s41598-019-54804-9

14. Thomas F, Rochaix P, White-Koning M, et al. Population pharmacokinetics of erlotinib and its pharmacokinetic/pharmacodynamic relationships in head and neck squamous cell carcinoma. Eur J Cancer. 2009;45(13):2316–2323. doi:10.1016/j.ejca.2009.05.007

15. Hopkins, AM, Menz, BD, Wiese, MD, et al. Nuances to precision dosing strategies of targeted cancer medicines. Pharmacol Res Perspect. 2020; 8:e00625. https://doi.org/10.1002/prp2.625

16. Gérard S, Bréchemier D, Lefort A, et al. Body composition and anti-neoplastic treatment in adult and older subjects – A systematic review. J Nutr Health Aging. 2016;20(8):878–888. doi:10.1007/s12603-015-0653-2

17. Wang Y, Jiang T, Qin Z, et al. HER2 exon 20 insertions in non-small-cell lung cancer are sensitive to the irreversible pan-HER receptor tyrosine kinase inhibitor pyrotinib. Ann Oncol. 2019;30(3):447–455. doi:https://doi.org/10.1093/annonc/mdy542

18. Gao X, Miao R, Tao Y, Chen X, Wan C, Jia R. Effect of Montmorillonite powder on intestinal mucosal barrier in children with abdominal Henoch-Schonlein purpura: A randomized controlled study. Medicine (Baltimore). 2018;97(39):e12577. doi:10.1097/MD.0000000000012577

19. Pérez-Gaxiola G, Cuello-García CA, Florez ID, Pérez-Pico VM. Smectite for acute infectious diarrhoea in children. Cochrane Database Syst Rev. 2018;4(4):CD011526. doi:10.1002/14651858.CD011526.pub2

20. Magnoli AP, Alonso VA, Cavaglieri LR, Dalcero AM, Chiacchiera SM. Effect of monogastric and ruminant gastrointestinal conditions on in vitro aflatoxin B1 adsorption ability by a montmorillonite. Food Addit Contam Part A Chem Anal Control Expo Risk Assess. 2013;30(4):743–749. doi:10.1080/19440049.2013.784398

21. Effinger A, O’Driscoll CM, McAllister M, Fotaki N. Impact of gastrointestinal disease states on oral drug absorption - implications for formulation design – a PEARRL review. J Pharm Pharmacol.2019;71(4):674–698. doi:10.1111/jphp.12928

22. Gunes A, Coskun U, Boruban C, et al. Inhibitory Effect of 5-Fluorouracil on Cytochrome P450 2C9 Activity in Cancer Patients. Basic Clin Pharmacol Toxicol. 2006;98(2):197–200. doi:10.1111/j.1742-7843.2006.pto_304.x

